# Coming out of the ashes we rise: Culturally and linguistically diverse international nursing students during the COVID-19 pandemic

**DOI:** 10.1101/2024.05.13.24307209

**Authors:** Eric Lim, Linda Ng, Huaqiong Zhou, Ambili Nair, Fatch Kalembo

**Affiliations:** Curtin University, School of Nursing Australia; The University of Southern Queensland, School of Nursing and Midwifery Australia

**Author notes:** **Contribution:** EL - Study conception and design, Data collection, Data analysis and interpretation, Drafting of the article; LN - Data collection, Drafting of the article, Critical revision of the article; HZ - Data collection, Critical revision of the article; AN - Data collection, Critical revision of the article; FK - Critical revision of the article. **Topic/question:** Qualitative study of culturally and lingusistically diverse international student who stayed in Australia during COVID-19 pandemic.

**Keywords:** Australia, international students, culturally and linguistically diverse, mental health, learning experiences, COVID-19, pandemic, resilience

## Abstract

**Background and aim:** Research on international students conducted during the COVID-19 pandemic has persistently highlighted the vulnerabilities and challenges that they experienced when staying in the host country to continue with their studies. The findings from such research can inevitably create a negative image of international students and their ability to respond to challenges during unprecedented times. Therefore, this paper took a different stance and reported on a qualitative study that explored culturally and linguistically diverse (CaLD) international nursing students who overcame the challenges brought about by the pandemic to continue with their studies in Australia.

**Method:** A descriptive qualitative research design guided by the processes of constructivist grounded theory was selected to ascertain insights from participants’ experiences of studying abroad in Australia during the COVID-19 pandemic.

**Results:** Three themes emerged from the collected data that described the participants’ lived experiences, and they were: 1) *Viewing international education as the pursuit of a better life*, 2) *Focusing on personal growth*, and 3) *Coming out of the ashes we rise*.

**Discussion:** The findings highlight the importance of recognising the investments and sacrifices that CaLD international students and their families make in pursuit of international tertiary education. The findings also underscore the importance of acknowledging the qualities that CaLD international students have to achieve self-growth and ultimately self-efficacy as they stay in the host country during a pandemic.

**Conclusion:** Future research should focus on identifying strategies that are useful for CaLD international nursing students to experience personal growth and ultimately self-efficacy and continue with their studies in the host country during times of uncertainty such as a pandemic.

## INTRODUCTION

All around the world, people are choosing to relocate to other countries for a wide range of reasons such as economic prospects, environmental conditions, personal safety, and employment (Forbes-Mewett, 2020). To increase their opportunities of finding employment, many are choosing to make substantial investments to pursue higher education overseas (Forbes-Mewett, 2020; Karram, 2013; Tallon et al., 2021). Prior to the COVID-19 pandemic, Australia used to attract international students from over 190 countries (Australian Government, 2021), with a large and growing proportion of culturally and linguistically diverse (CaLD) students coming from non-western countries (Khawaja et al., 2014). Culturally and linguistically diverse (CaLD) refers to a population that encompasses a variety of cultural backgrounds and languages. This term is often used to describe groups of people who come from different ethnic, linguistic, or national origins within a particular society or community. It highlights the diversity present within a population in terms of cultural practices, beliefs, and languages spoken (Pham, et al., 2021). The top-ranking non-western countries in descending order included India, China, Malaysia, Indonesia, Sri Lanka, the Republic of Korea, the Hong Kong Special Administrative Region of the Republic of China, and Singapore (Guo, 2010; Hawthorne, 2014; Lim et al., 2016).

Most of the CaLD international students in Australia are young and unseasoned travellers who are leaving their homes for the first time to live alone in a foreign country (Australian Government, 2012). As such, they may be making significant lifestyle adjustments and need to acculturate to the differing social-cultural differences while living and studying in a foreign environment (Wang et al., 2008). Previous findings highlighted that CaLD international students were more conservative, introverted, and less ready to express their views with other people due to their cultural backgrounds, traditions, and family influences (Yan & Berlinder, 2013). The differences between the culture of origin and the culture of contact can make them feel out of place and powerless in the new environment – Cultural shock (Yan & Berlinder, 2013). Consequently, CaLD international students are more susceptible to poor mental health and negative learning experiences when compared to domestic students, especially if they struggle with language barriers, isolation, and acculturation stress, (King et al., 2020; Mitchell et al., 2017; Vardaman & Mastel-Smith, 2016).

As such, it is important to ensure that their learning experiences are positive, supportive, and satisfying (Lim et al., 2023; Tallon et al., 2021).

### Background

COVID-19 is a global pandemic that has impacted all sectors of the community including universities and the education of health sciences students (Agu et al., 2021). The international higher education sector was one of the first to be significantly impacted because of border closures and travel restrictions (Sidhu et al., 2021). Although many CaLD international students moved back to their country of origin (Van de Velde et al., 2021), a huge number remained in Australia despite the comment made by the former Australian Prime Minister, “at times like this, if you are a visitor to this country, it’s time to make your way home” (Gallagher et al., 2020 pg.1).

At the start of the pandemic, CaLD international students who stayed in Australia to carry on with their studies were not eligible for emergency relief measures introduced by the government (Hari et al., 2021; Nguyen & Balakrishnan, 2020). As such, many of them had to work longer hours to support their studies, maintain food and housing security, and reduce the financial burden placed on their families (Hari et al., 2021). For instance, Asian international students had to increase their working hours which placed additional stress on their ability to uphold their academic standards (King et al., 2020). Individuals whose jobs were affected due to lockdowns experienced higher levels of stress if they struggled to find new employment quickly to meet or keep up with their expenses (Van de Velde et al., 2021).

At the peak of the pandemic, many of the CaLD international students, specifically those from Chinese backgrounds were allegedly experiencing an increase in racial discrimination from members of the community (King et al., 2020). The experience of racism can negatively impact their sense of belonging and self-esteem, contributing to poor mental health (King et al., 2020). To add to their distress, CaLD international students had limited support from their families and friends due to travel restrictions and lockdowns, with communication only possible through phone or online means (Aslan & Pekince, 2020; King et al., 2020). The loneliness and physical distancing from loved ones exacerbated their level of stress, anxiety, and sadness, and if poorly managed could lead to long-term psychological impairments (Aslan & Pekince, 2020).

In addition, the pandemic disrupted normal learning and teaching patterns, with face-to-face classes transitioning to online formats and changes in examination and assessment methods (Agu et al., 2021; Van de Velde et al., 2021). As such CaLD international students needed to make sure that they had adequate technology and internet services, learn how to navigate online learning platforms, and adapt to the changes in their learning environments to maintain a positive learning experience (Agu et al., 2021; Van de Velde et al., 2021). The majority of the CaLD international students come from middle-class backgrounds, where their families often make significant financial sacrifices to support their education abroad. In some cultures, underperforming academically can evoke deep feelings of shame and guilt for these students, as they perceive it as letting down their families (King et al., 2020).

### Aim of the study

Literature highlights that CaLD international students who chose to stay in their host countries during the pandemic showed admirable resilience and adaptability (Beckstein, 2020). As such, the aim of this study was to explore the lived experiences of CaLD international nursing students who decided to remain in Australia to continue with their studies at two Australian universities during the COVID-19 pandemic. The research question that guided this study was, *‘What are the lived experiences of culturally and linguistically diverse (CaLD) international nursing students who chose to remain in Australia for their studies amidst the COVID-19 pandemic?’*

#### Ethical considerations

The study proposal and participant information sheet were sent together with the email to participants who contacted the researchers listed on the flyers of every campus and university.

Participants were provided an explanation about the study and that participation in the study was voluntary and would not affect their learning at the university. Participants were also assured that the chosen interview mode (either face to face or online) was anonymous and confidential.

Participants were advised that they could withdraw anytime during the interviews and if they withdraw, their data will not be used for the study. During the interview, consent was obtained verbally if the interview was online and digitally if face to face. Ethical approval was obtained from Curtin University Human Research Ethics Office (HRE2022-0238) and The University of Southern Queensland Ethical Review Commitee (H22REA114).

## METHODS

### Design

A descriptive qualitative study that followed the processes as outlined in the constructivist grounded theory (Charmaz, 2006) was chosen to allow the researchers to gain insight into the participants’ experiences of studying abroad in Australia during the COVID-19 pandemic (Tenny et al., 2023). The use of this study design enabled the researchers to follow a rigorous process to obtain an in-depth exploration and develop an understanding of the individuals’ lived experiences (Dodgson, 2017).

### Participants

We decided to employ the purposive sampling technique as it enables us to concentrate on specific groups of individuals who can offer insightful and pertinent data, effectively addressing the research aim of this study (Nyimbili, & Nyimbili, 2024). CaLD international nursing students who meet the eligibility criteria of (i) 18 years or older and (ii) have the lived experience of staying in Australia during the Covid-19 pandemic (2019 – 2021) were invited to participate in the study. These students were either in the 2^nd^ year or 3^rd^ year or had recently graduated from Australia at the time of participation. Their shared lived experiences allowed the researchers to gain profound and insightful insights into the participants’ encounters.

### Settings

This study took place post-pandemic restrictions where teaching and learning activities were gradually returning to the on-campus mode of delivery. As such, the researchers were able to recruit participants from the school of nursing of [removed for peer-review] and [removed for peer-review] through an advertisement made on the school’s webpage, blackboard announcements, flyers on-campuses, and personalised emails sent to all students who were identified as onshore international students.

### Recruitment

The CaLD international nursing students who were interested in the study contacted the researchers of their respective universities and were provided with the participant information sheet, informed consent form, and interview guide prior to setting a date for the interview.

Participants had the option of being interviewed face-to-face or virtually via Microsoft Teams at their convenience.

### Data collection

All the participants provided their informed consent and completed a basic demographic questionnaire prior to the commencement of the interview. The semi-structured interview method was chosen as it allows reciprocity between the interviewer and participant, enabling the interviewer to improvise follow-up questions based on the participant’s responses (Kallio, et al., 2016). The questions were informed broadly by literature (Naz, et al., 2022) around the themes of financial support, family support, social support, work, and housing. The semi-structured interviews were conducted independently by four members of the research team (Authors 1 to 4) who were experienced in conducting qualitative, research. All the interviews were conducted using an interview schedule between the period of October 2022 to April 2023. Face-to-face interviews were recorded digitally and transcribed verbatim using Microsoft Word, and virtual interviews were video recorded and transcribed using Microsoft Teams. Initial coding of the transcriptions was performed by Author 1 after each interview, and this process ensured that data collection was guided by data saturation, whereby no new information was collected in subsequent interviews (Kyngäs, 2020).

### Data analysis

Rigour is the means of demonstrating the plausibility, credibility, and integrity of the qualitative research process. According to Kenny, et al (2024), rigour of a study can be determined if actions and developments of the researcher can be examined. This study utilises the framework proposed by Guba and Lincoln (1981) for assessing the rigour of qualitative research: credibility (field notes, recording), transferability (thematic logs), auditability (field notes, recording and thematic logs) and confirmability (audit trail by constant comparison) (Lincoln & Guba, 1985).

In this study, all the interviews were digitally recorded with the permission of the participants. The audio recording ensured that an identical replication of the contents of each interview was available to facilitate analysis. Audio recording was chosen as a tool for record keeping because it could greatly increase the quality of field observations (Giles, et al., 2024) by allowing the researcher to analyse, interpret, and report the participants’ own words. The audio recording can be used to refute criticism that qualitative research is prone to systematic bias (Tuckett, 2005).

The importance of ensuring that interview transcripts are verbatim accounts for what took place is widely accepted (Eftekhari, 2024). A verbatim transcript is a faithful reproduction of an aural record, taken as an unquestionable record of the interview and as an expression of truth (Eftekhari, 2024).

Field notes refer to various notes recorded by the researcher during or after her observation of a specific phenomenon. Tuckett (2005) defines field notes as descriptions of experiences and observations the researcher has made while participating in an intense and involved manner. These brief notes allowed the investigators to improve the discussion, by keeping track of ideas and themes, coming back to them for clarification and further discussion and elaboration (Tuckett, 2005).

The initial codes were compared and grouped by Author 1 using the constant comparison analysis technique central to the grounded theory (Charmaz, 2006). This process ensured that the codes were grouped and categorised according to their meanings. The developing categories were checked by all members of the research team and any discrepancies found in the constructed categories were reviewed and discussed until consensus was achieved. This peer review process eliminated the potential biases in Author 1’s interpretations of the data, and this ensured the trustworthiness of the findings (Holloway & Galvin, 2016).

Next, the sentences and segments that were grouped under each of the developing categories were re-read and scrutinised for themes and concepts (Charmaz, 2006). This process allowed meanings of the categories to emerge and were grounded in the lived experiences of the participants. The emerged meanings of the categories were then related to the literature to ascertain the degree of support for the ideas generated (Kolb, 2012; Tucket, 2005). To attain dependability, this study followed the COREQ Checklist for Interviews and Focus Groups (Tong et al., 2007) to write up the findings.

## RESULTS

We adjusted our recruitment methods to include verbal advertisements during lectures, tutorials, and simulation laboratory sessions. Despite utilising numerous methods of recruitment (flyers, community hubs, study desks, verbal during lectures, tutorials, and simulation laboratory sessions) at various campuses, only twenty CaLD international students responded. Arrangements were made with these twenty students, however only 19 students agreed to participate. The recorded interviews ranged from 30 to 60 minutes in duration with a total of 10 hours and 04 minutes. Eight of the participants chose to be interviewed in person, and the rest were virtually interviewed. The CaLD international nursing students who participated in this study consisted of 17 females and two males. Eleven participants were aged between 20 to 29, seven were aged between 30 to 39, and one was aged between 40 to 49. As shown in Table 1, the majority of the CaLD were from China (36.8%), followed by Malaysia (21.1%), Nepal (15.8%), Hong Kong (5.3%), India (5.3%), Kuwait (5.3%), Singapore (5.3%), and South Korea (5.3%).

**Table 1.** Demographic data of participants.

| Participant number | Country of origin | Gender | Age group | Relationship status | Family and relatives' status | Employment status | Highest qualification | Length of stay in Australia |
| --- | --- | --- | --- | --- | --- | --- | --- | --- |
| 1 | Malaysia | Female | 20 to 29 | In a relationship | Family/relatives here: Yes<br>Lives with family/relatives: Yes<br>Carer responsibility: No | Paid work: Yes<br>1 – 20hrs a week | High school | More than 4 years |
| 2 | Hong Kong | Female | 20 to 29 | In a relationship | Family/relatives here: No<br>Lives with family/relative: No<br>Carer responsibility: No | Paid work: Yes<br>1 – 20hrs a week | Undergraduate | More than 4 years |
| 3 | China | Female | 20 to 29 | In a relationship | Family/relatives here: Yes<br>Lives with family/relative: No<br>Carer responsibility: No | Paid work: Yes<br>1 – 20hrs a week | Undergraduate | More than 4 years |
| 4 | China | Female | 30 to 39 | Single | Family/relatives here: Yes<br>Lives with family/relative: No<br>Carer responsibility: No | Paid work: Yes<br>1 – 20hrs a week | Undergraduate | More than 4 years |
| 5 | Malaysia | Male | 20 to 29 | Single | Family/relatives here: No<br>Lives with family/relative: No<br>Carer responsibility: No | Paid work: Yes<br>More than 20hrs a week | Undergraduate | More than 4 years |
| 6 | China | Female | 30 to 39 | Married | Family/relatives here: Yes<br>Lives with family/relative: Yes<br>Carer responsibility: Yes | Paid work: Yes<br>1 – 20hrs a week | Postgraduate | More than 4 years |
| 7 | Kuwait | Female | 40 to 49 | Married | Family/relatives here: No<br>Lives with family/relative: No<br>Carer responsibility: Yes | Paid work: Yes<br>More than 20hrs a week | Postgraduate | Less than 1 year |
| 8 | Singapore | Male | 20 to 29 | Single | Family/relatives here: No<br>Lives with family/relative: No<br>Carer responsibility: No | Paid work: Yes<br>More than 20hrs a week | Undergraduate | 3 to 4 years |
| 9 | Nepal | Female | 30 to 39 | Single | Family/relatives here: No<br>Lives with family/relative: No<br>Carer responsibility: Yes | Paid work: Yes<br>1 – 20hrs a week | Undergraduate | More than 4 years |
| 10 | China | Female | 30 to 39 | Married | Family/relatives here: Yes<br>Lives with family/relative: Yes<br>Carer responsibility: Yes | Paid work: Yes<br>1 – 20hrs a week | Postgraduate | More than 4 years |
| 11 | China | Female | 30 to 39 | Married | Family/relatives here: Yes<br>Lives with family/relative: Yes<br>Carer responsibility: Yes | Paid work: Yes<br>More than 20hrs a week | Vocational training | More than 4 years |
| 12 | India | Female | 20 to 29 | Single | Family/relatives here: Yes<br>Lives with family/relative: Yes<br>Carer responsibility: No | Paid work: Yes<br>1 – 20hrs a week | Undergraduate | 3 to 4 years |
| 13 | China | Female | 30 to 39 | In a relationship | Family/relatives here: No<br>Lives with family/relative: No<br>Carer responsibility: No | Paid work: Yes<br>1 – 20hrs a week | Undergraduate | More than 4 years |
| 14 | China | Female | 30 to 39 | Married | Family/relatives here: Yes<br>Lives with family/relative: No<br>Carer responsibility: No | Paid work: Yes<br>1 – 20hrs a week | Undergraduate | More than 4 years |
| 15 | Malaysia | Female | 20 to 29 | Single | Family/relatives here: No<br>Lives with family/relative: No<br>Carer responsibility: No | Paid work: Yes<br>More than 20hrs a week | High school | Less than 1 year |
| 16 | Malaysia | Female | 20 to 29 | Single | Family/relatives here: No | Paid work: Yes | High school | 1 to 2 years |
|  |  |  |  |  | • Lives with family/relative: No<br>• Carer responsibility: No | • 1 – 20hrs a week |  |  |
| 17 | Nepal | Female | 20 to 29 | Single | • Family/relatives here: No<br>• Lives with family/relative: No<br>• Carer responsibility: No | • Paid work: Yes<br>• 1 – 20hrs a week | • High school | • 1 to 2 years |
| 18 | Nepal | Female | 20 to 29 | Married | • Family/relatives here: No<br>• Lives with family/relative: No<br>• Carer responsibility: No | • Paid work: Yes<br>• 1 – 20hrs a week | • High school | • 1 to 2 years |
| 19 | South Korea | Female | 20 to 29 | Single | • Family/relatives here: No<br>• Lives with family/relative: No<br>• Carer responsibility: No | • Paid work: No | • High school | • Less than 1 year |

The majority of the CaLD international nursing students who participated in this study reported that they had lived in Australia for more than 3 years (68.4%) at the time of data collection. The highest qualifications of the participants were: high school (n = 6), vocational training (n = 1), undergraduate (n = 9), and postgraduate (n = 3). Close to three-quarters of the participants (73.7%) did not live with their family or relatives, 63.2% did not have family or relatives living in Australia, and 94.7% of the participants were engaged in paid work

### The experiences of CALD international nursing students during the Covid-19 pandemic

There was a consensus among the participants that their decisions to remain in Australia were inevitably met with harsh adversaries. Three themes emerged during the process of data analysis that represented the reasons and experiences of CaLD international nursing students. The themes were: (i) *viewing international education as the pursuit of a better life*, (ii) *focusing on personal growth, and* (iii) *coming out of the ashes we rise* (see Figure 1. Coding tree).

**Figure 1.**
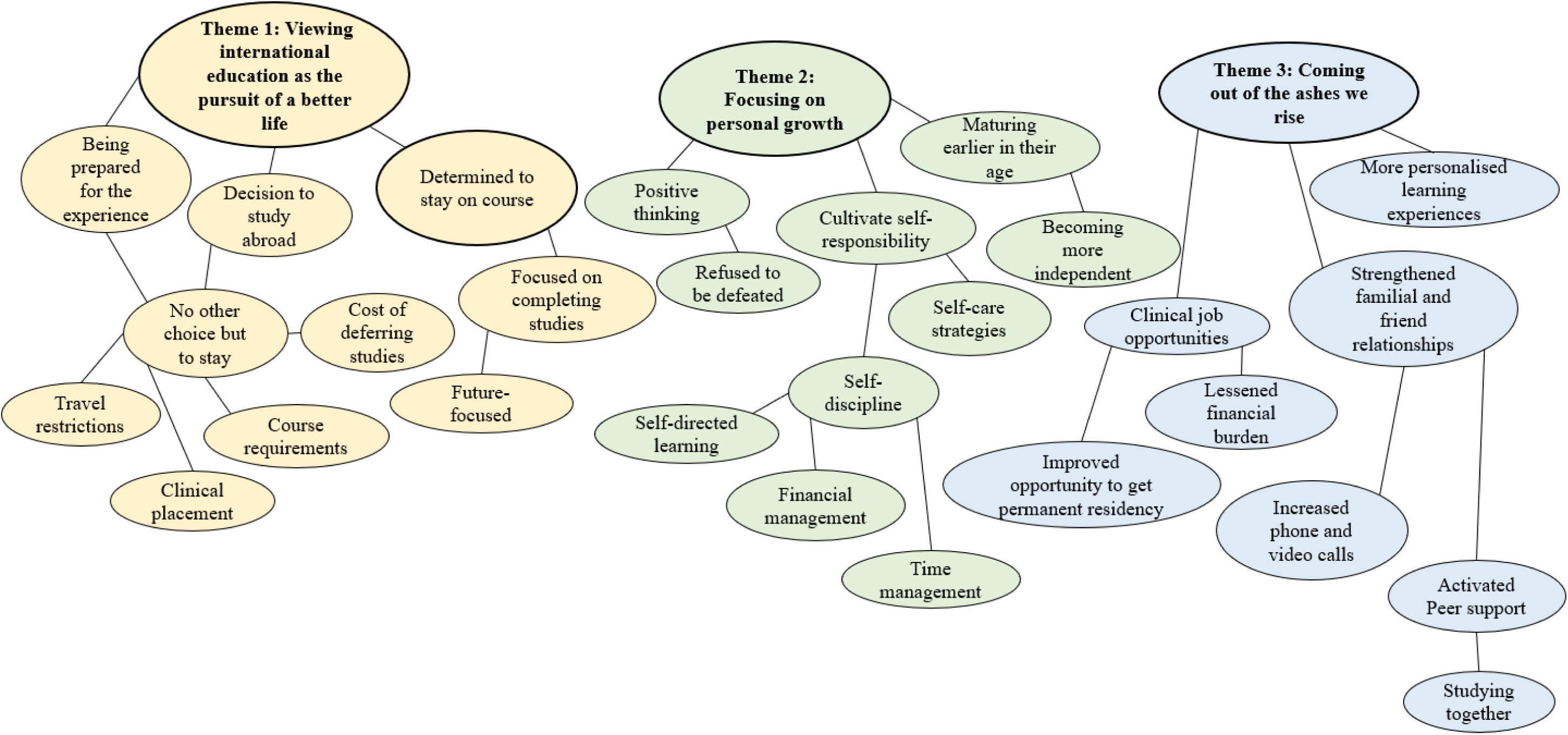
Coding tree.

### Theme 1: Viewing international education as the pursuit of a better life

The first theme *‘viewing international education as the pursuit of a better life’* represented the mindset of many of the CaLD international nursing students who participated in this study. There was a consensus among the participants that studying overseas was viewed as a major life-changing decision that included making significant investments and sacrifices including spending an extended period away from their family or loved ones. Two of the participants stated: *“It’s a big money, so I don’t wanna give up, so just keeping going on.”* (P13); *“Everything changed but what can I do? I just have to accept it* (P17)”. For this reason, the majority of the participants shared that they were determined to remain in Australia to complete their nursing course despite having the knowledge of the impact of living in a pandemic: *“I’m pursuing a career in nursing so that’s why I need to stay and keep studying”* (P10); as “*nursing contains clinical placement and the only way to pass the course is to attend the clinical placement. If I go back to China and I can’t come back, I will have to extend my studies.”* (P3); “*I haven’t finished my degree so I have to stay here”* (P5); *“I’m just afraid that if I go back to my home country, I couldn’t return to Australia to continue my study”* (P2). Interestingly, most of the participants revealed that their knowledge of the travel restrictions imposed on students was not particularly distressing for them. As one of the participants said:

> *“There’s no choice… and if I went back home, it was the same condition, and the situation would be worse if I went back because I had to study online from there. So basically, having no choice kind of motivates me because I know that I’ll have to deal with it” (P12)*.

Nevertheless, a few of the participants highlighted that they were caught by surprise by the prolonged period of travel restrictions: *“I came from Kuwait and was aware of the travel restriction. But there were chances that the international border will reopen and they [social media] were giving us hope that it might be opened within one to two months”* (P6); *“Honestly, I thought that it was only for a short while, I didn’t know that there would be a lockdown for three years. I only thought that it would be only for three months”* (P4). Those who were caught by surprise by the prolonged restrictions on international travel were reportedly more likely to experience homesickness: *“I didn’t know that it’s gonna be three years… I’ll be able to go home if I miss home… COVID having that option taken away during the pandemic”* (P4). Yet, there was a consensus among the participants including those who were homesick that they would still choose to remain in Australia to continue with their studies in pursuit of a better life: *“In hindsight, I know that even if there is a lockdown. I would still come, I would still fly here, despite the risk of not coming back. I will still fly and try to get before a lockdown”* (P4); *“Yes, I will come as I am very much career focused”* (P6).

### Theme 2: Focusing on personal growth

The second theme *‘focusing on personal growth’* described the attitudes of CaLD international nursing students who participated in this study. Many of the participants highlighted the importance of accepting their decision to remain in Australia to reduce the negative impacts on their mental health and wellbeing. One of the participants declared: *“I convinced myself that it is the best decision… whatever is going to happen will happen, so I have no control over that and just take it positively”* (P9). Other participants shared how they engaged in positive thinking to maintain their mental health and wellbeing: *“There’s less restriction here compared to China. Lots of people that go back ended up having anxiety or depression”* (P3); *“The Australian government did a really good job about COVID when compared with my home country. If I go back to China, I’m not sure if I can get used to the restrictions there” (P10)*; *“I think the learning environment and healthcare are much better in here compared with my home country during COVID”* (P11). As one of the participants stated:

> *“I’m happy that they didn’t make us leave Australia. Some students who signed up to go to Melbourne during the pandemic had to literally finish their degree in Malaysia, but they have paid for coming to Aussie. So, I’m happy I’m able to study in Australia to get the life experience that they don’t have.” (P4)*

Many of the participants who were aged between 20 to 29 highlighted that they *“felt that they had to mature earlier than their peers [who are living in their country of origin] of the same age”* (P1), and *“learn to cope as this is the first time, they experienced something like COVID”* (P6). The participants perceived the need to cope with challenges like COVID-19 independently, without burdening their families. As one of the participants stated:

> *“We have decided to stay in Australia and the finances are way higher than what we like to have back home… we know that we have to pay fees and all [other expenses] … So that’s all on us and is our responsibility to pay for our fees… we cannot blame others but ourselves… we have to keep telling ourselves that it is all on us [because we made that decision] and then work on it, rather than [to rely on family or loved ones]” (P9)*.

For this reason, most of the CaLD international nursing students who participated in this study highlighted that they learned to juggle their studies and paid work. This helped them to acquire skills to manage their time and finances more effectively. One of the participants shared:

> *“I have to manage all the time and money, to work and study so that I don’t struggle. I have to work and save up enough for me to pay my next semester’s tuition fee, pay my rent, and pay for my food. I don’t spend on entertainment … I believe that is what most international students’ life would be [remaining in Australia during the COVID-19 pandemic” (P11)*.

Another participant stated:

> *“It is definitely the experience you never expected to have back home. I didn’t even cook all my life. I didn’t look after myself like my family used to do everything for myself. I just used to do whatever I wanted to… No responsibilities or anything. Nothing more of that now… Being here by yourself, looking after yourself, paying rent. Arranging finances and all, that’s the experiences [maturing earlier than their peers of the same age] I was talking about” (P9)*.

The need for CaLD international nursing students to assume responsibility for their own lives also motivated them to change their lifestyles, live healthier, and learn coping strategies to strengthen their psychological, emotional, and physical wellbeing: *“I did a lot of mediation… and focused on those things [which are important]. I think it is good because I didn’t have time to think and worry about other problems”* (P14); *“I had a friend who was very depressed, but we motivated him to play sports. He started playing cricket, and he is much better now”* (P6). One of the participants added:

> *“I changed myself a lot [personal growth] during the COVID because I know how important health is. So, before I went to the university campus to study, I never used to go to the gym but now I start attending some of the gym classes. I met some of the people there and did some exercise with them… I guess I made some friends from the gym… before that, I don’t have any friends in here” (P3)*.

### Theme 3: Coming out of the ashes we rise

The last theme *‘Coming out of the ashes we rise’* commemorated the achievements of the CaLD international nursing students who participated in this study. Interestingly, when asked to reflect on their experiences, most of the participants claimed to have had a positive experience of living in Australia during the COVID-19 pandemic: *“It helped me build some resilience. I’ve finished my studies and managed to get a job as a nurse”* (P12); *“Hundred percent! I will still choose to stay here if COVID returns. I learned so much about myself that I will never get to if I went home”* (P1).

Most of the participants highlighted that they were pleased about the abundance of clinical job opportunities for international nursing students, which was not the case prior to the COVID-19 pandemic. As such, they were able to lessen the financial burdens placed on themselves and their families or loved ones: *“During COVID, there were lots of chances to get work, especially for nursing… I work for the agency in a casual position like personal care assistant”* (P3); *“I managed to work as an AIN while studying throughout the pandemic”* (P4); *“I managed to support my mother and sister [financially] who were stuck here with me due the travel restriction”* (P1). Some of the participants were able to secure ongoing paid work in the healthcare sector and this has helped them to qualify for getting their permanent residency in Australia. As one of the participants stated:

> *“If I am not studying nursing, I probably cannot find a job during this situation. You know now it’s easier than before for nursing students to get a job, so we are very lucky, and I think the best choice. If I didn’t choose to stay in Australia during COVID, my husband and I cannot get the PR [Permanent Residency] invitations” (P7)*.

Many of the participants also claimed that they felt less stressed with the changes to the teaching patterns as they were able to personalize their learning experiences with their lifestyles. One of the participants stated:

> *“Our studies were moved online but I like it. I don’t have to wake up early to attend lectures or go to tutorials. I just signed into Zoom and the lectures were recorded so it works well for me. Then our exams were also online which to me was good because I can do the exam at home, and I don’t get stressed while traveling to the campus to sit the exam” (P15)*.

Another participant who shared the same sentiment said: *“I was just happy that I have a break. I was already exhausted with placements and assessments, so I was happy. I get to sleep in and no need to wake up early”* (P16). Additionally, many of the participants shared that their relationships with their family, loved ones, and friends were improved as there was stronger communal support and understanding:

> *“We talk to each other, work with each other. If one of us got COVID, the other one would go out to buy groceries, so I depend heavily on housemates and close friends from my own country… we kind of understand each other that we are out of the country, and we don’t have anyone else here helping us” (P5)*.

There was also a stronger peer support to support one another with their studies: *“I have a friend from church, and she organized an English corner for the international students to study together”* (P10). Those participants who have family or loved ones here in Australia found that they have more time to spend with them and the increased engagement has supported them to feel closer. As one of the participants explained: “*My partner is here, and we have to face all the struggles together, do things together and that it just makes me feel that life is more interesting with him”* (P2). On the other hand, those participants who do not have family or loved ones here in Australia found themselves *“spending more time catching up with close friends and family in [their home country] online or phone calls”* (P11). All of these highlights how CaLD international nursing students overcame the challenges brought about by the COVID-19 pandemic and emerged victoriously on the other side.

## DISCUSSION

In line with previous research on international students during the COVID-19 pandemic, CaLD international nursing students in this study encountered similar challenges (Gallagher et al., 2020; Gomes et al., 2021; Nguyen & Balakrishnan, 2020). Most of the studies conducted on international students during COVID-19 have continually highlighted their poor mental health and wellbeing. Consequently, CaLD international students were often perceived as lacking in resilience to remain in the host country during the pandemic (Gallagher et al., 2020; Gomes et al., 2021; Nguyen & Balakrishnan, 2020).

Yet, the findings in our study revealed opposing evidence of CaLD international students achieving positive lived experiences while staying in Australia amidst the border closures and travel restrictions. More specifically, the findings in our study highlighted that international students were motivated and determined to stay in the host country to continue with their studies, thus reinforced the importance of having temporary relief support financial assistance to support international students to remain in Australia (Ramia, et al., 2022). It underscored the importance for policymakers and decision-makers in education to rethink their approach to the pandemic, seeing it as a chance to foster a sense of belonging among international students. This entails nurturing their personal development and empowering them to pursue independent living, rather than simply advising them to go back to their home countries.

The findings of our study highlighted the qualities of CaLD international students such as their maturity, determination, and adaptability when pursuing international education. These attributes hold immense importance for the academic progression of international students, especially during challenging times as noted by Sabouripour and Roslan (2015). However, they often remain unnoticed amidst the more common hurdles like acculturation stress and language barriers encountered by CaLD international students. There is a need for more research that focuses on identifying the strengths and capabilities of CaLD international students to remain in the host country during the pandemic. The growing body of research findings has the potential to influence policy adjustments aimed at enhancing support for international students to stay in their host countries. Education policymakers and decision-makers must acknowledge the financial commitments and sacrifices made by CaLD international students and their families to pursue education abroad. Our study underscores that the majority of CaLD international students opt for overseas study in search of improved opportunities for a better life. The lack of humanistic regard for CaLD international students during the pandemic underscored the urgent need for more inclusive and empathetic support systems (Van de Velde et al., 2021).

CaLD international students and their families who felt neglected, abandoned, and devalued by the Australian Government during the pandemic (Van de Velde et al., 2021), may perceive themselves as mere commodities or ‘cash cows’ to the Australian economy (Soong & Maheepala, 2023). This can discourage future CaLD international students from choosing Australia as their host country for international education, thus significantly impacting the total education industry’s revenue and employment (Thatcher, et al., 2020). It was reported in a recent survey of Indian international students that they preferred the US or Canada over Australia or New Zealand post-pandemic (Freeman, et al., 2022).

### Significance of findings to nursing education and practice

The findings from our study provided valuable insights into the lived experiences of CaLD international students who continued to stay in Australia to continue with their nursing program during the COVID-19 pandemic. The results of our study showed that CaLD international nursing students who participated in this study displayed more strengths, resilience, and maturity to continue with studies onshore when compared to other CaLD international students in the other programs, for example, business, mining, or engineering. This highlighted that international students should not be treated as a homogenous group during the COVID-19 pandemic (Australian Government, 2012).

The results of our study also implied the need for education policy- and decision-makers to acknowledge the uniqueness of international students from different backgrounds and programs that they were studying when implementing strategies to reduce the impacts on the country during unprecedented times. For example, nursing students, both local and international, were considered essential workers during the COVID-19 pandemic to address the critical nursing workforce shortages. However, there was a lack of foresight to fund nursing programs nationally and provide support for international nursing students to continue with their studies. Nevertheless, the findings of our study showed that CaLD international nursing students were able to tap into the increased paid work opportunities (Gómez-Moreno, et al., 2022) to achieve financial independence (Dempsey, et al., 2023).

Lastly, our findings showed that the majority of the CaLD international nursing students were juggling between their work and studies and welcomed the changes to normal learning and teaching patterns during the COVID-19 pandemic. This finding highlighted the need to personalise the nursing curriculum to improve the learning experiences (Middleton & Moroney, 2019), perhaps by incorporating the use of more modern technology to overcome the cost, time, and logistic problems (Hirt & Beer, 2020).

### Limitations of this study

This study is not without its limitations. First, the participants in this study consisted of CaLD international nursing students who were predominately from Asian countries. Therefore, the findings generated in this study may not be representative of the whole population of international nursing students who remained in Australia during the COVID-19 pandemic. Nonetheless, the study provides us with some indication of which students may be most vulnerable in a pandemic. Second, we only investigated the international nursing students from two regional universities in Australia so the findings may differ among other universities, areas, or populations such as students belonging to other disciplines/programs.

## CONCLUSION

This study explored the lived experiences of CaLD international nursing students who stayed in Australia during the COVID-19 pandemic to continue with their studies. The findings of this study provided important insights into the qualities that CaLD international students possessed to overcome the challenges brought about by the pandemic and emerge victoriously. Future research will be conducted to identify strategies that CaLD international nursing students identified as useful for them to activate personal growth and ultimately self-efficacy to stay and continue with their studies during times of uncertainty.

## RELEVANCE TO CLINICAL PRACTICE

The findings of our study highlighted the importance of education policy- and decision-makers to acknowledge the investments and sacrifices that CaLD international students and their families made to pursue international education and identify more humanistic approaches to support them to stay in Australia during unprecedented times. The findings of this study also highlighted the need to reconceptualise times of uncertainty such as a pandemic as opportunities for CaLD international students to experience self-growth, achieve self-efficacy, and ultimately actualise their talents and potentialities, which is the true purpose and goal for them to pursue international education (Shkoler & Rabenu, 2023).

## Data Availability

All data produced in the present study are available upon reasonable request to the authors

## Notes

### Competing Interest Statement

The authors have declared no competing interest.

### Funding Statement

This study did not receive any funding

### Author Declarations

Ethical approval was obtained from Curtin University Human Research Ethics Office (HRE2022-0238) and The University of Southern Queensland Ethical Review Committee (H22REA114).

